# Versus Arthritis Musculoskeletal Disorders Research Advisory Group Priority Setting Exercise Protocol

**DOI:** 10.1101/2021.10.04.21264485

**Authors:** Z Paskins, F Manning, C Farmer, C Le Maitre, E Clark, D Mason, C Wilkinson, D Andersson, T Barlow, F Bishop, C Brown, A Clark, D Dulake, M Gulati, R Jones, J Loughlin, M McCarron, N Millar, H Pandit, G Peat, S Richardson, E Salt, J Taylor-Wormald, L Troeberg, R Wilcox, E Wise, S Rudkin, FE Watt

## Abstract

Involving research users in setting priorities for musculoskeletal research is essential to raise awareness of the unmet needs for MSK research, to ensure research outcomes are patient-centred and relevant, have a high likelihood of resulting in patient benefit, reduce research waste and increase research value and impact. In 2018, Versus Arthritis convened an MSK Disorders Research Advisory Group (RAG) which included people with arthritis, health care professionals and researchers in MSK, in order to identify and prioritise research areas with a long-term aim of improving quality and impact of MSK research. On further review, there were few previous prioritisation approaches in this area looking across discovery science to more clinical research, at important research questions which might be common to a range of disorders or approaches incorporating input at all stages of the process by a range of stakeholders including people with arthritis. The group identified that more work to define research priorities in these areas was justified and designed a research priority setting process for MSK disorders. This manuscript documents the methodology that was developed by the group for this process.

**Methods:** Following a review, the Child Health and Nutrition Research Initiative (CHNRI) method for research prioritisation was selected as best aligning with the needs of this process. The group agreed on adaptations to the CHNRI approach, context, purpose and remit of the exercise and identified through consensus four priority research Domains: Mechanisms of disease; Diagnosis (including early diagnosis) and measuring the impact of these disorders; Living well with MSK disorders and Successful Translation. From all published CHNRI scoring criteria for generated research avenues or themes of research, the group identified six which were most relevant to this process. To ensure accessibility of the survey and scoring, these were refined to three: Equity (considered cross cutting, not scored but considered throughout the process), Importance (Will research in this area have potential to lead to important new knowledge) and Impact (Might research in this area make a difference). Importance and Impact were to be scored on a scale of 1-10 for each research avenue with equal weighting of these two criteria in the subsequent generation of a total score.

**Data collection:** Following ethical approval, an electronic first survey asking for important research uncertainties in the four research domains and any other areas will be distributed to all stakeholders (people with arthritis, researchers in all stages of MSK disorders research, healthcare professionals, industry e.g. pharmaceutical and medical technology companies, research funders, healthcare providers, government policy makers and charities). The next step is to consolidate all the gathered research uncertainties from the first survey into finalised research domains and avenues. Uncertainties will be summarised using deductive thematic analysis and organised into possible themes which will then be considered and refined by each of four appointed subgroups within the RAG. Following group and lay review and refinement of the wording including tests of readability, the second survey including this full list of research avenues will be submitted for ethical approval. The second survey will be completed by the same range of stakeholders as the first survey, both those who previously completed and new respondents. Respondents will be invited to rate each research avenue using the two scoring criteria, with the avenues presented in a random sequence to avoid bias.

**Analysis Plan:** All available data will be analysed, from all respondents completing the survey in full and all partial respondents. For each research avenue, a mean criterion score will be calculated for each of the two criteria from all available survey responses (considering the number of respondents in each case), and then the two mean criterion scores will be summed to create a total score. Response rates and missing data for scoring of avenues will be reported. The primary prioritisation output of this exercise will be to produce a single ranked list of these total scores of research avenues, from highest to lowest. The most highly ranked avenues will be highlighted, for example the top five to top ten overall and from each research domain, with the exact number and nature of this depending on the distribution of the data. Respondent characteristics will be summarised including self-identified stakeholder group, age group, gender and ethnic background, to describe the diversity and representation within the survey respondents as far as possible.

**Dissemination plan:** Findings will be communicated in a number of formats, both written and spoken, to ensure accessibility to all stakeholders, and will also be used by the charity in internal strategy development. Dissemination will include the submission of a manuscript to a peer-reviewed journal.

## 1. Introduction: setting Priorities for Musculoskeletal Disorders Research

Musculoskeletal (MSK) disorders include more than 150 disorders that affect the musculoskeletal system, including osteoarthritis and back pain. These conditions are characterised by pain and impaired physical function, often increasing the risk of immobility, obesity, other chronic physical and mental health conditions and all-cause mortality [Briggs et al, 2018]. MSK disorders affect an estimated 18.8 million people across the UK, with one in five people consulting annually for a MSK disorder [Versus Arthritis, 2019]. In the UK, MSK disorders alone account for more than 22% of the total burden of ill health and account for the third largest area of NHS programme spending at £4.7 billion, with a substantial cost including joint replacement and other forms of surgery [Versus Arthritis, 2019]. Furthermore, the impact on societal costs is great. MSK disorders are the leading cause of work disability, sickness absence from work, ‘presenteeism’ and loss of productivity across Europe, resulting in lost productivity as high as 2% of gross domestic product [Bevan, 2015].

MSK disorders can be inflammatory (such as rheumatoid arthritis) or non-inflammatory. As the risk of many non-inflammatory MSK disorders increases with age, and the UK population profile is becoming older, the prevalence of these disorders is set to increase. This association with age has also led to the normalisation, tolerance and de-prioritisation of these conditions and the pain and loss of function that result from them. Despite the prevalence and impact of MSK disorders, people living with these conditions report impaired quality of life, reduced independence and not getting the support that they need [Versus Arthritis, 2019]. Research into MSK disorders has been demonstrated to produce a 25% economic return on investment - considerably greater than the rate of return for cancer research (10%) and the UK government minimum threshold for investment (3.5%) [Versus Arthritis, 2019; Glover et al, 2014]. Yet, when compared with other disorders such as cancer, funding for research into MSK disorders is disproportionately low, compared to the burden of disease [Versus Arthritis, 2019]. Furthermore, within funding for MSK disorders, research into non-inflammatory disorders is disproportionately low, compared to funding for inflammatory disorders, in view of the high prevalence of many of these conditions. In recognition of the huge impact of these disorders, prevention, early detection and treatment of MSK disorders across the life course has recently become a Public Health England priority [PHE, 2019].

Involving research users (such as clinicians, patients, academics, policy makers) in setting priorities for research is essential to ensure research outcomes are patient-centred and relevant, have a high likelihood of resulting in patient benefit, reduce research waste and increase research value and impact [Chalmers et al, 2014]. Previous exercises to set priorities in musculoskeletal research have either focused on individual musculoskeletal disorders [Sheehan et al, 2019], or focused on a range of disorders but not used formal prioritisation methods [EULAR, 2019]. This study aims to fill this gap by conducting a formal research prioritisation setting exercise across non-inflammatory musculoskeletal disorders and throughout all stages of research.

## 2. Methods

### 2.1. Advisory group and context

In 2018, Versus Arthritis convened an MSK Disorders Research Advisory Group (RAG) in order to identify and prioritise research areas with a long-term aim of improving quality and impact of MSK research. The group meets at least three times a year and comprises 26 members including people with arthritis (PwA), researchers, and healthcare professionals, facilitated by the charity. Among clinical and research members, there is wide representation of clinical specialties (physiotherapists, GPs, rheumatologists, orthopaedic surgeons), in disease interest and research stage (discovery scientists, epidemiologists, clinical and applied health services researchers). Public contributors (in this case, PwA) bring experience of a range of MSK conditions. The group is committed to diversity, with a balanced mix of gender, BAME representation, four nations representation and stage of career/age.

Early activities of the group included defining our population of interest: people with MSK disorders, osteoarthritis, crystal diseases such as gout, primary and secondary causes of musculoskeletal pain including regional and widespread pain (such as back pain, shoulder pain and tendinopathy, other regional pain syndromes and fibromyalgia), hypermobility, metabolic bone disorders (such as osteoporosis and rare diseases) and musculoskeletal injuries caused by acute traumatic events.

In July 2019, the Group met at a face-to-face meeting and carried out an exercise to define the current landscape within research into MSK disorders, identifying strengths, weaknesses, opportunities and threats. Like the group, it considered all types and stages of research, thinking across disorders and disciplines. The output from this exercise was summarised in an internal report. Four main themes were identified by the group as areas of important potential impact and/or research need. These were:

- Mechanisms of disease;
- Diagnosis (including early diagnosis) and measuring the impact of these disorders;
- Living well with MSK disorders (including clinical management and implementation);
- Successful translation.

These four themes or domains are shortened within this protocol to Mechanisms; Diagnosis; Living Well and Translation.

On further review of all available existing research prioritisation exercises in this area by the group (such as Rheumamap, James Lind Alliance Priority Setting Partnerships, prior Versus Arthritis Clinical Studies Group priorities), it was noted that there were few approaches looking across discovery science to more clinical research, at important research questions which might be common to a range of disorders or approaches incorporating input at all stages of the process by a range of stakeholders including PwA. The group recognised that more work to define research priorities in these areas was therefore justified. The RAG appointed a group lead for each of these four themes, each with expertise in the relevant area. A PwA was appointed to lead the group ‘Living well with MSK Disorders’.

### 2.2. Overview of method and rationale

The group activities thus identified a need for a further research priority setting exercise in this area. The group wished to review and select an appropriate research prioritisation method aligned to their needs. The group agreed that the selected method needed to be:

- valid (used by others, current)
- incorporating a range of stakeholder perspectives,
- combining both discovery science, clinical and applied health services research questions,
- incorporating objective scoring.

Following a review of methods including the James Lind Alliance, Delphi, the Combined Approach Matrix, the Essential National Health Research method and Child Health and Nutrition Research Initiative (CHNRI) method [Yoshida, 2016], the CHNRI [Rudan et al, 2008] was selected as best aligning with the needs of the group.

The original CHNRI approach comprises four main steps, overseen by an advisory group:

i. gather research questions or uncertainties
ii. consolidate these uncertainties
iii. score the consolidated uncertainties
iv. analyse for prioritisation, taking into account stakeholder views.

A modified approach was proposed, which had been used by others [Shah et al, 2016], to ensure all stakeholders and research users are involved at each stage, rather than just in the final stage. The CHNRI uses specific terms to catalogue suggested areas of research as shown in Table 1 [Rudan et al, 2008]. Research avenues are scored and prioritised. We adopted this terminology.

**Table 1:** CHNRI terminology used in this study

|  |  |
| --- | --- |
| Research Domain | Broad areas of research, corresponding to the four themes we identified within MSK |
| Research Avenue | More specific area of research within a domain |
| Research Options | More specific area of research, within an avenue, corresponding to 3-5 year programme of research |
| Research Questions | More specific questions within a research option, corresponding to a single research paper |

Subsequently the group agreed on adaptations to the CHNRI approach, agreed context, purpose and remit of the exercise (Table 2) and specified four priority research domains: mechanisms of disease; diagnosis and impact; managing and living well with MSK disorders; and, successful translation, informed by their earlier work as discussed. It was also agreed that important questions which were not covered by these headings should also be collected. The group agreed criteria by which to score research questions, based on accepted CHNRI approaches. These were subsequently refined following lay review.

**Table 2:** Context, purpose and remit of MSK research priority setting exercise

|  |  |
| --- | --- |
| Motivation | MSK disorders affect an estimated 18.8 million people across the UK, with one in five people consulting their GP annually for an MSK disorder. In the UK, MSK disorders account for more than 22% of the total burden of ill health and contribute substantially to the problem of multi-morbidity. MSK disorders account for the third largest area of NHS programme spending, after mental health and disorders of circulation, at £4.7 billion.<br>However, despite the high prevalence, and the personal, social and economic impact of these conditions, the needs of people with MSK disorders are not being met; MSK disorders are often normalised and not prioritised. There are no research priority setting exercises which have considered MSK disorders collectively and crossed all research stages to identify shared priorities. |
| Long-term goal | To improve the quality and impact of research which seeks to develop our understanding and management of MSK conditions.<br>• to enhance prevention, early detection and treatment and care of MSK disorders<br>• to improve the quality of life and wellbeing of those with MSK disorders<br>• to reduce personal, social and economic burden of MSK disorders |
| Population of interest | People (aged 18 or over) with, or at risk of MSK disorders, their families, carers and healthcare providers.<br>MSK disorders defined as osteoarthritis, crystal disease such as gout, primary and secondary causes of musculoskeletal pain including regional and widespread pain (such as back pain, shoulder pain and tendinopathy, other regional pain syndromes and fibromyalgia), hypermobility, metabolic bone disorders (such as osteoporosis and rare diseases) and musculoskeletal injuries caused by acute traumatic events.<br>Participants and research questions within the United Kingdom (UK) were identified as the main focus, although the findings may well have relevance beyond the UK. |
| Timeframe for impact | Both within, and beyond 3 years (0-3 years (short term), >3 years (longer term)) |
| Research domains identified a priori | • Mechanisms of disease<br>• Diagnosis and impact<br>• Living well with MSK disorders<br>• Successful translation<br>Low and high-risk research will be considered. |
| Audience | • People with arthritis<br>• Researchers of all stages<br>• Healthcare professionals<br>• Funders and other agencies who influence the funding and research strategy of musculoskeletal conditions<br>• Industry, such as pharmaceutical and medical technology companies |

Research prioritisation exercises that rely on surveys alone to elicit research questions or uncertainties are potentially subject to risk of missing important research questions, as a result of response bias [Rudan et al, 2017]. For this reason, the advisory group considered whether to include existing research prioritisation exercises within the ‘gathering’ stage. It was decided that existing research prioritisation exercises should not be included, because: many of these previous priorities are disease specific with under-representation of certain disorders; their inclusion could lead to bias and dilute new insights; it was not felt acceptable to score priorities that have already undergone a scoring or selection process; and, inclusion would increase the size of the second survey, threatening completion rates. Instead, it was agreed the finalised priorities garnered through this process would be compared and contrasted with existing priorities in the discussion of the final report.

### 2.3. Step 1: Gathering uncertainties

Following appropriate ethical approval, an electronic first survey asking for important research uncertainties will be distributed to all stakeholders including people with arthritis, researchers, healthcare professionals, industry e.g. pharmaceutical and medical technology companies, research funders, healthcare providers, government policy makers and charities using Versus Arthritis existing mailing lists, social media, and professional networks primarily accessed by the group.

The survey will be written by the group. Those completing survey 1 will be asked to comment on four priority domains of musculoskeletal disorder research previously identified by the RAG. Respondents may provide responses to all four domains or select areas according to their interests. Respondents will also have the opportunity to highlight any other uncertainties in a ‘catch-all’ section of survey. In particular, respondents will be asked about “important unanswered questions or uncertainties”, including their opinion on shared priorities across different disorders, disciplines and stages of research.

Participants will be asked to provide optional information on their demographics and background including details of employer, research experience and interests (if relevant), to give confidence that the characteristics of survey responders are understood. Where there are low numbers of respondents with particular characteristics whilst the survey is open, we will adjust recruitment strategies to try to compensate accordingly. Respondents will be given the option to consent to future contact for a second scoring survey (see below). The first survey will be open for 4-6 weeks, depending on completion rates.

### 2.4. Step 2: Consolidation of uncertainties

The objective of this step is to consolidate all the gathered research uncertainties from the first survey into finalised research domains and avenues, the latter of which will undergo scoring. The process of consolidation of research uncertainties is summarised in Figure 1.

**Figure 1:**
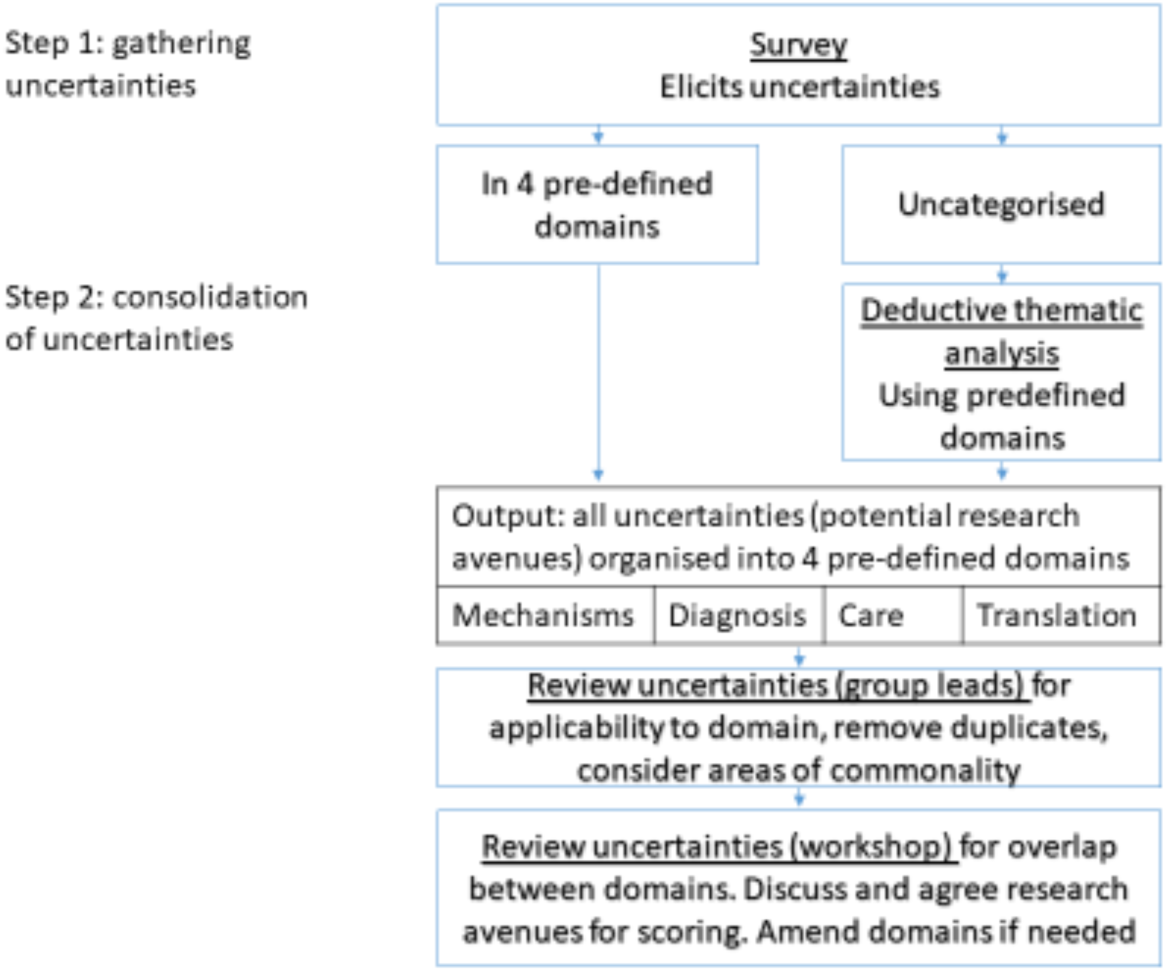
Process of gathering and consolidating research uncertainties

Respondent data from survey 1 will be handled and summarised by Versus Arthritis (the data controller). Identifiable and demographic information will only be handled by Versus Arthritis. GDPR and data protection requirements will be adhered to at all times. Information about data storage and use by Versus Arthritis and e-consent for this will be sought from respondents as part of the survey. Demographic information will be held and summarised separately to the uncertainties/questions in the survey data to protect the identity of individuals. This information will be used to describe the demographics and background of the overall population responding to the survey, to ensure diversity and the level of representation of different groups. Only summarised data will be made available to researchers and no contact details or personal/identifiable data will be shared.

Group members will view and work with the survey 1 response data on a shared restricted Microsoft Teams site hosted by Versus Arthritis. All members of the group handling or viewing the data will first agree to a memorandum of understanding relating to its use, as required by Versus Arthritis as the data controller.

Uncertainties from the survey will be collected in the four pre-defined research Domains (see Table 2; ‘Mechanisms’, ‘Diagnosis and impact’, ‘Living well’ and ‘Translation’) and in an un-categorised ‘Other’ section. Initially, a researcher with experience of thematic analysis and understanding of both discovery science and clinical research will conduct the following steps

1. Familiarisation with the data and data organisation; this will include preparing an Excel spreadsheet with
  a. one uncertainty per line (multiple uncertainties will be separated)
  b. numbering uncertainties, to enable linkage back to the respondent number, comment number and in some cases, the original domain
  c. Repeating context where needed (indicated in {}) from original entries when multiple uncertainties are separated, to allow interpretation
2. Identifying any uncertainties which are
  a. potentially unclear and may need further interpretation – identifying if there is any relevant context in other uncertainties entered by the same participant which can aid understanding (again then added in in {})
  b. potentially in the wrong category. Two-three group members will discuss categorisation and interpretation of membership of research Domains where necessary.
  c. Out of scope because comment is
    i. About children
    ii. About inflammatory arthritis
    iii. About auto-immune disorders
  These disorders are considered by other research advisory groups who will set their own priorities.
3. Conducting deductive thematic analysis to categorise any un-categorised uncertainties from the survey into one of the four pre-defined research Domains.
4. Creating a new analysis spreadsheet with reorganised uncertainties under appropriate Domains
  a. Organise into possible themes within each Domain for Domain/sub-group leads to consider and refine if necessary, which may inform generation of draft research avenues

When grouped into the four pre-defined Domains, lists of uncertainties will be reviewed at dedicated sub-group meetings by the respective Domain sub-groups, all including public contributors to i) ensure the uncertainties are applicable to their domain, ii) consider interpretation of any statements which are unclear and iii) review, edit and agree themes and iv) start to draft potential research Avenues based on these agreed themes, ideally starting with a verb e.g. evaluate, determine, investigate.

At all stages, a clear audit trail of decision-making will be kept, including which original uncertainties have informed which research avenues. Maintenance of link from research Domain and research Avenue to source questions will be ensured, to ensure full transparency and evidencing of the output.

A 1–2-day workshop of the RAG will be convened. The Domain leads will present draft research Avenues within their Domains to the whole group. The group will consider the appropriateness of the original four domain areas. Following agreement of domain areas, the RAG will divide into ∼four working groups relating to the identified domains to refine the wording of research Avenues, with attention to consistency across the Domains and balance between Avenues which are not too broad or specific. These will be presented to and agreed by the whole group through iterative discussion. Readability measures will be deployed, aiming for as low a reading age as feasible.

The research Avenues and accompanying survey information composed by the group (second survey) will then be reviewed by external lay review, refined and submitted as an ethical amendment, prior to advertising.

### 2.5. Step 3: Scoring research questions

The RAG reviewed a range of possible criteria by which to score each research Avenue as part of methods development, using small-group working and then whole-group consensus. From the criteria previously associated with the CHNRI method [Rudan, 2008, 2014], the group felt answerability, deliverability, equity, feasibility, potential impact on disease burden, novelty and potential for translation were the most applicable across the 4 domains. Feasibility was considered important but a more challenging domain to score by non-experts in the field, so not suited to all intended survey respondents. The group initially suggested three criteria which they felt incorporated all of these areas (impact, importance and feasibility) but on further external lay review considered that two criteria were sufficient to provide essential information for ranking, whilst minimising the length and burden of the second survey and maximising accessibility to all. The applicable criteria were therefore refined into two broad criteria, relating to impact and importance. In addition, *Equity* was considered a cross-cutting criterion that should be reflected throughout the process and criteria, but not specifically scored. For example, in disease burden, could the research address, or worsen, inequalities in health.

Each broad criterion was defined by one scoring question with a supporting explanatory statement.

A second survey including questions to score each research avenue against these criteria will be developed by the MSK RAG, including public contributors within and out with the group. This electronic survey will be distributed to participants from Step 1 who gave their consent to further contact as well as being advertised more broadly, through similar channels to the first survey. Balance of participation of different groups will be monitored whilst the survey is live, particularly promoting participation of low responding groups from the first survey. RAG members will be excluded from participating in the second survey. Questions will be presented to respondents in a random order, within each of the four Domains (again presented in a random order), to enable data from partial respondents to be used. The anonymised characteristics of respondents providing this information will be reviewed to ensure diversity in terms of all predefined characteristics (i.e., to avoid over or under representation of any one particular group). For this second survey, this will include optional additional questions on age, sex and ethnicity, as well as similar questions for the first survey (except for their preference on research Domain or any request for identity/email).

Respondents in the second survey will be asked to score to what extent each research Avenue meets each of the two criteria, using a numeric rating scale (1-10). Such a continuous scale was felt important to use in methods development by the group, rather than a binary (Yes/No) scale used in some CHNRI exercises. Options including a 1-5 scale were explored but 1-10 option was favoured by the group. An option to answer the question ‘I am unsure’ will also be available. Answers can also be missed. An option to save and return will be available. It will be made clear that each respondent should answer based on their own knowledge and perspective.

This list of research Avenues will be presented in the survey for scoring (for each of the two criteria) with 2 pre-agreed ‘reminder’ stems for the criteria (abbreviated from Table 3):

**Table 3.** Selected CHNRI scoring criteria for the exercise

| Criterion | Question | Explanation |
| --- | --- | --- |
|  | These statements will be in column headers on each page of the survey. | These notes will be listed at the top of each page of the survey. |
| Importance | Will research in this area have potential to lead to important new knowledge?<br>(Where 1 is not likely and 10 is extremely likely) | It might help to consider<br>• Will this lead to significant new important knowledge or understanding, **at any stage of research?**<br>• What is already known or being studied in this research area? |
| Impact | Might research in this area make a difference?<br>(Where 1 is not likely and 10 is extremely likely) | It might help to consider<br>• Could this research lead to new or more effective treatment or tests?<br>• How much could this research result in patient benefit, whether in the short or long term?<br>• Consider the size of this impact, (you might score a small benefit for a large number of people, or a large benefit for a smaller number of people)? |

- Will this research lead to important new knowledge?
- Will this research make a difference and lead to impact?

Both surveys will be piloted prior to going live, within the group and by external volunteers. Again, respondent data from Survey 2 will be handled and summarised separately to the rest of the survey data, by Versus Arthritis, to report the demographics of those responding. Only summarised data will be accessible to researchers and no contact details or personal/identifiable data.

### 2.6. Step 4: Analysis and prioritisation

Summary: For each research avenue, a ‘mean criterion score’ will be calculated for each of the two criteria, and then the two mean criterion scores will be summed to create a ‘total score’. The primary prioritisation output of this exercise will be to produce a ranked list of these total scores, irrespective of the research Domain.

Detail of the primary reporting: All available data will be analysed, from all respondents completing the survey in full and all partial respondents.

Response rate: The exact timing window of the survey, the total number of people consenting to the survey, the number of complete and partial responders will be reported. In supplementary data, for each criterion of each Avenue, the total number of responses will be reported for all respondents, for relevant stakeholder groups and the proportion of missing data given.

Calculation of means: The sum of each of the impact and importance criterion scores for each research avenue will be calculated (individual mean criterion scores will also be reported as supplementary data, for all participants, and for stakeholder groups e.g. patients, healthcare professionals and researchers, and complete and partial responders). The mean criterion score for impact and for importance will be calculated separately, as these may have different numbers of respondents. An appropriate measure of dispersion will also be given.

For each research avenue:

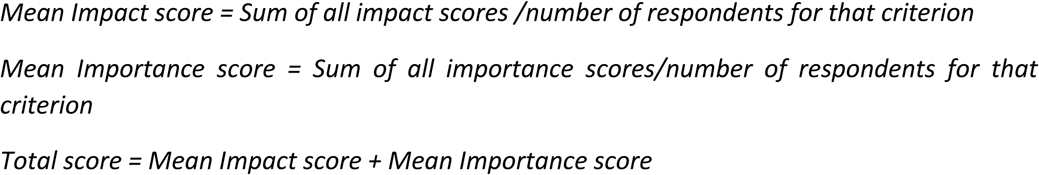

i.e., The denominator for the mean calculation will always be the total number of responders (excluding those who scored ‘Unsure’) to that particular question, meaning that non-response or, being ‘unsure’ will not adversely affect scoring. (This CHNRI method using mean scores assumes that the distribution of responses approaches a normal distribution: this will be verified, with appropriate methods modification if this is not the case).

The group discussed weighting criteria differently, but chose to support equal weighting of the two criteria and presenting a single ranked ordering summarising the findings.

Prioritisation: All research Avenues will then be presented in a single rank order, providing their total scores, from highest to lowest. The most highly ranked Avenues will be highlighted, for example the top five to top ten overall and from each research Domain, with the exact number and nature of this depending on the distribution of the data. This is typical for a CHNRI process.

Respondent characteristics will be summarised including self-identified stakeholder group (and for researchers, the type of research they conduct), age group, gender and ethnic background, to describe the diversity and representation within the survey respondents as far as possible. N.B. these respondent characteristics will not be available for those who partially complete the survey because these questions are at the end of the survey.

### 2.7. Step 5: Dissemination

Findings will be communicated in a number of formats, both written and spoken, to ensure accessibility to all stakeholders, and will also be used by the charity in internal strategy development. Dissemination will include the submission of a manuscript to a peer-reviewed journal.

The publication of results will include the pre-defined methodology of this process and the breakdown of criterion scores within each research Avenue in case of relevance to different audiences.

The results will be used to inform Versus Arthritis research strategy and this will be disseminated by the charity to various stakeholders in a number of formats, including website, public reports and government submissions.

### 2.8. Ethical approval and data collection

All public facing surveys will be approved via a University Research Ethics Committee (Medical Sciences Division Oxford). All amendments (including the text for Survey 2) will similarly be approved, prior to use. This approval allows for the storage and analysis of data for this purpose and the publication of summary data from this exercise.

Appropriate online survey tools will be used, as permitted by Versus Arthritis as the data controller. For Survey 1, MS Forms is envisaged. For Survey 2, the use of Smart Survey is envisaged (for additional functionality to allow randomisation of questions). All survey data will be held and summarised by Versus Arthritis.

### 2.9. Timeline

Step 1 October –November 2020

Step 2 December to March 2021

Survey 1 closes 15^th^ Jan 2021

15^th^ Jan until 28^th^ Feb 2021 - Survey results reviewed, thematic analysis

1 – 31st March 2021 – generation of draft and then refined research avenues by subgroups

April – July 2021 – lay review and refinement of avenues and draft survey; ethical approval of Survey 2 text including final list of research avenues for scoring

Step 3 August - October 2021

Survey 2 closes 4^th^ October 2021

Step 4 October - November 2021

Step 5 December 2021 onwards

The changed ways of working brought about by the Covid pandemic may affect some of these timelines.

## Data Availability

These data will not be publicly available or available to others due to the nature of the consent and restrictions of data collection and storage (Versus Arthritis is the data controller). Summary data from the priority setting exercise will be available on reasonable request to Versus Arthritis.

## 4. Appendix

**Appendix Table 1:** Version Control of protocol

| Version Number | Date | Comments |
| --- | --- | --- |
| 2.0 | 02.10.21 | Finalised ahead of analysis |
| 1.2 | 28.9.21 | More detail added on analysis plan<br>Scoring criteria reduced from 3 to 2 |
| 1.1 | 30.6.21 | Amendments to description of scoring following lay review |
| 1.0 | 13.1.21 |  |

**Appendix Table 2:**
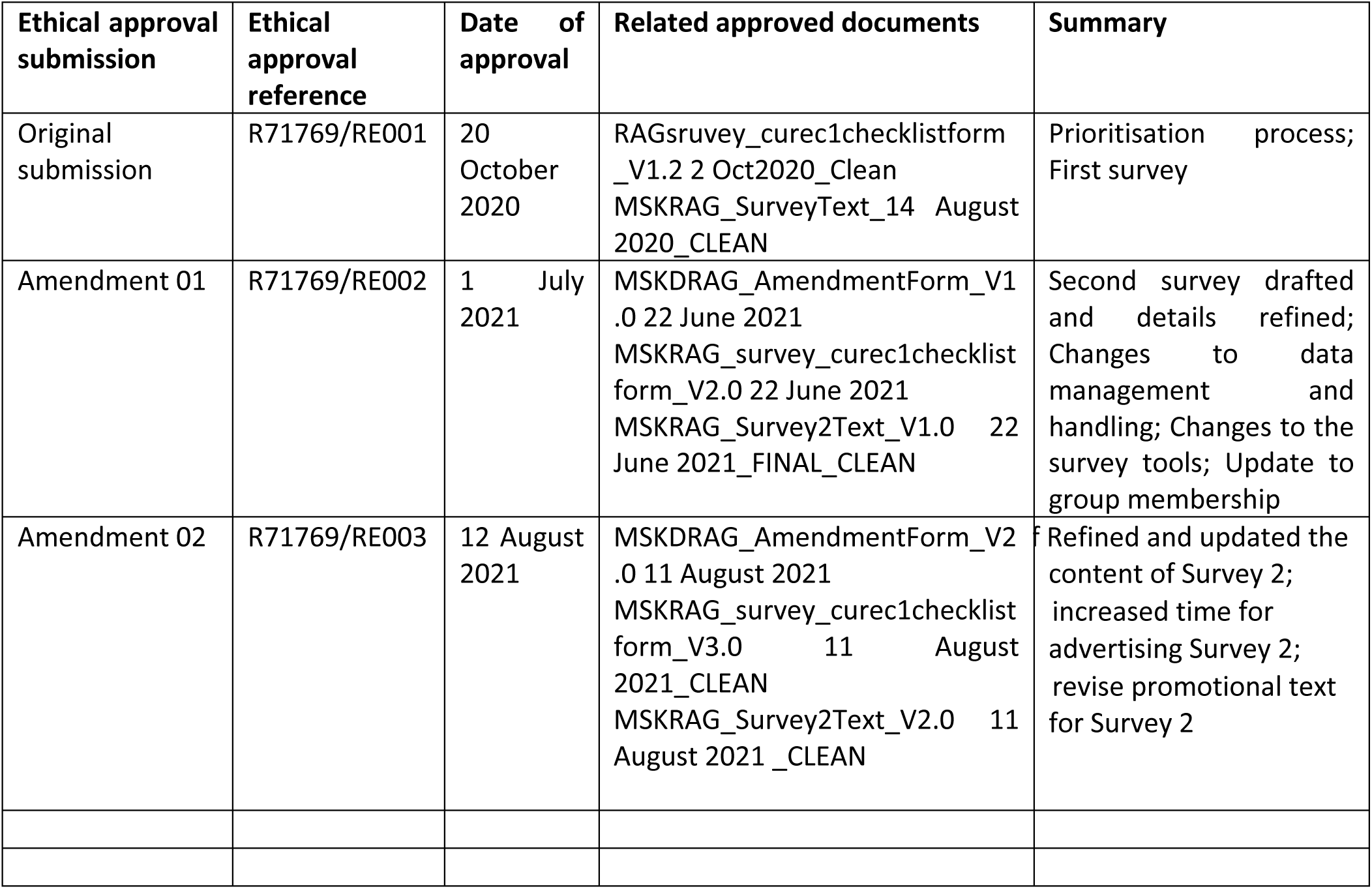
Ethical approvals and related other documents

| Ethical approval submission | Ethical approval reference | Date of approval | Related approved documents | Summary |
| --- | --- | --- | --- | --- |
| Original submission | R71769/RE001 | 20 October 2020 | RAGsurvey_curec1checklistform_V1.2 2 Oct2020_Clean<br>MSKRAG_SurveyText_14 August 2020_CLEAN | Prioritisation process;<br>First survey |
| Amendment 01 | R71769/RE002 | 1 July 2021 | MSKDRAG_AmendmentForm_V1 .0 22 June 2021<br>MSKRAG_survey_curec1checklist form_V2.0 22 June 2021<br>MSKRAG_Survey2Text_V1.0 22 June 2021_FINAL_CLEAN | Second survey drafted and details refined; Changes to data management and handling; Changes to the survey tools; Update to group membership |
| Amendment 02 | R71769/RE003 | 12 August 2021 | MSKDRAG_AmendmentForm_V2 .0 11 August 2021<br>MSKRAG_survey_curec1checklist form_V3.0 11 August 2021_CLEAN<br>MSKRAG_Survey2Text_V2.0 11 August 2021_CLEAN | Refined and updated the content of Survey 2; increased time for advertising Survey 2; revise promotional text for Survey 2 |

